# An open annotated dataset and baseline machine learning model for segmentation of vertebrae with metastatic bone lesions from CT

**DOI:** 10.1101/2024.10.14.24314447

**Authors:** Nazim Haouchine, David B. Hackney, Steve D. Pieper, William M. Wells, Malika Sanhinova, Tracy A. Balboni, Alexander Spektor, Mai A. Huynh, David E. Kozono, Patrick Doyle, Ron N. Alkalay

**Affiliations:** Beth Israel Deaconess Medical Center, Boston, MA; Brigham & Women’s Hospital, Boston, MA; Harvard Medical School, Boston, MA; Isomics, Inc., Cambridge, MA

## Abstract

Automatic analysis of pathologic vertebrae from computed tomography (CT) scans could significantly improve the diagnostic management of patients with metastatic spine disease. We provide the first publicly available annotated imaging dataset of cancerous CT spines to help develop artificial intelligence frameworks for automatic vertebrae segmentation and classification. This collection contains a dataset of 55 CT scans collected on patients with various types of primary cancers at two different institutions. In addition to raw images, data include manual segmentations and contours, vertebral level labeling, vertebral lesion-type classifications, and patient demographic details. Our automated segmentation model uses nnU-Net, a freely available open-source framework for deep learning in healthcare imaging, and is made publicly available. This data will facilitate the development and validation of models for predicting the mechanical response to loading and the resulting risk of fractures and spinal deformity.

## Introduction

After the liver and the lungs, bones are the third most frequent target site of metastases in the United States (1), with the spine being the most frequent site of bone metastases (2). Up to 50% of spinal metastases receive treatment, and 5-10% receive surgical management (3). Patients are at high risk for skeletal adverse events, including bone pain, fracture, hypercalcemia, and neurological dysfunction secondary to the spinal cord or cauda equina compression (4), that profoundly and negatively impact patients’ lives (24), requiring high doses of narcotics and reduced quality of life (5). Although the impact of spinal metastasis on the vertebral column anatomical integrity is readily imaged with MRI and CT (6,7,8), in the absence of quantitative assessment (9, 10, 11), standard-of-care imaging modalities (12, 13, 14) do not provide precise estimates of the structural competence of the spine. This limited ability profoundly negatively impacts patients’ clinical care, with clinicians forced to decide without objective quantitative criteria whether surgery or vertebral augmentation should be offered or withheld in attempts to treat to reduce the risk of fracture or to control pain. This choice can be particularly tragic when treatment could have been offered before the development of complications.

Implementing routine quantitative measurements from clinical imaging, such as vertebral segmentation, biomechanical quantification (26), lesion classification, longitudinal lesion growth monitoring (25), and risk progression estimates, would guide individualized earlier intervention. Recently, deep learning has been successfully applied to vertebral segmentation and classification tasks, demonstrating high accuracy and outperforming conventional methods (15). This success is due, in part, to the Large Scale Vertebrae Segmentation Challenge (VerSe) (16) and dataset (26), containing a total of 374 multi-detector CT scans with voxel-level annotations publically released. Before VerSe, only small public imaging datasets with vertebral segmentations existed (17,18,19,20,21,22), hindering efforts to develop robust and accurate segmentation and classification methods. Nevertheless, one limitation of VerSe is the exclusion of scans with vertebral abnormalities, such as bone metastasis and primary bone tumors, from their dataset.

In this work, we provide the first publicly available imaging dataset of vertebral segmentation masks for spine CT images of subjects with metastatic spine disease with annotations of vertebral lesions per vertebral level. This dataset is aimed to allow the development of automated large-scale deep learning methods for spine segmentation, vertebral level, and lesion classification and to facilitate longitudinal studies for monitoring lesion growth and predicting fracture risk.

## Materials and Methods

### Patient Overview

This dataset contains 55 metastatic spine disease patients CT scans (Male: Female 36:19; mean age: 64.70, range: 33-82, see Table 1) treated with radiotherapy or Stereotactic Body Radiation Therapy between September 2020 and October 2021 due to documented spine metastases (CT, MR, or bone scan imaging).

**Table 1:**
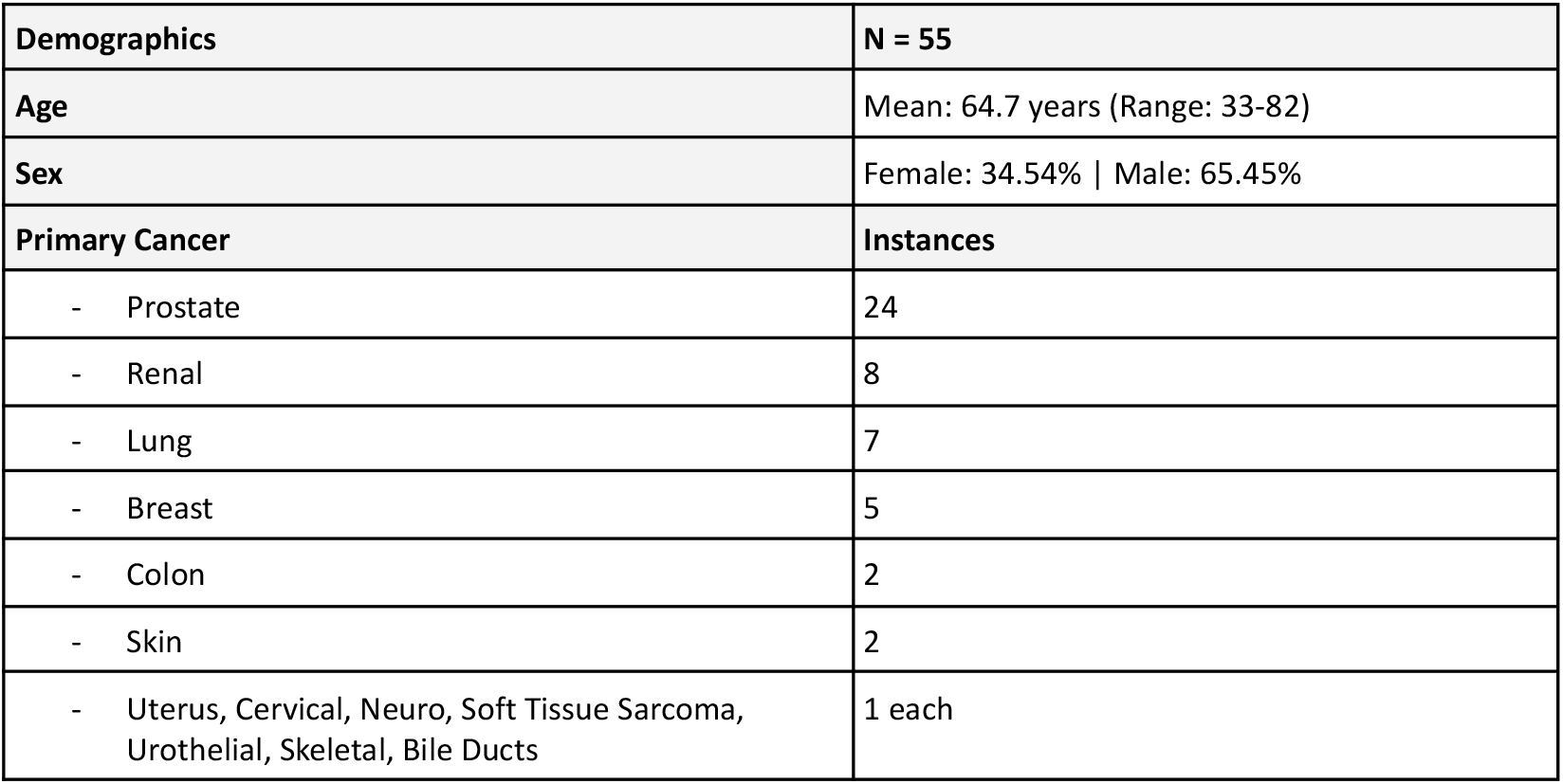
Patient demographics and primary cancer categories.

| Demographics | N = 55 |
| --- | --- |
| Age | Mean: 64.7 years (Range: 33-82) |
| Sex | Female: 34.54% \| Male: 65.45% |
| Primary Cancer | Instances |
| - Prostate | 24 |
| - Renal | 8 |
| - Lung | 7 |
| - Breast | 5 |
| - Colon | 2 |
| - Skin | 2 |
| - Uterus, Cervical, Neuro, Soft Tissue Sarcoma, Urothelial, Skeletal, Bile Ducts | 1 each |

Patients were identified for the study by the attending clinician according to the following inclusion criteria: (1) Histologically or cytologically documented stage IV bone metastases and radiographic (CT scan or bone scan) evidence of bone metastases, and (2) Karnofsky Performance Status >70. Patients were excluded if they had abnormal bone metabolism Paget disease, and untreated hyperthyroidism, hyperprolactinemia, or Cushing disease, or had previous surgery or vertebral augmentation at the site of radiation. Patients with prior radiotherapy treatment were excluded only if the patient had prior radiotherapy treatment at the spinal region under consideration. All contributions to this study were based on approval by Institutional Review Boards and were conducted following the 1964 Declaration of Helsinki.

### Image Acquisition

Clinical CT data was obtained on a General Electric scanner (LightSpeed RT16) or a Siemens scanner (SOMATOM Confidence). While acquisitions vary by patient, CT imaging was typically performed with an in-plane resolution of 0.3125×0.3125mm, in-plane matrix of 512×512, and slice thickness of 1.0-1.5mm. Data were fully de-identified with scans including no anatomy above the neck. All data were visually inspected before release. With the de-identified patients selected retrospectively, the study did not require informed consent from individual participants.

### Vertebral Segmentation, Level Annotation, and Lesion Classification

The CT data was imported into the 3DSlicer imaging platform. For each spine, eighteen vertebrae (T1 to L5) were considered for annotation depending on the field of view, with labels from 6 to 25 in the original segmentation label maps. Note that partially visible vertebrae at the top or bottom of the scan (or both) were not annotated. Each vertebral level was segmented using “Grow from Seeds” method, resulting in volumetric label maps by a spine biomechanics expert (RNA 25 years of experience). The anatomical accuracy of the segmentation and leabling were confirmed by an expert radiologist (DBH, 35 years of experience). In addition to the delineations, DBH and RNA classified vertebral metastatic lesions as Not-involved, Lytic, Blastic, or Mixed, as illustrated in Figure 2.

**Figure 1:**
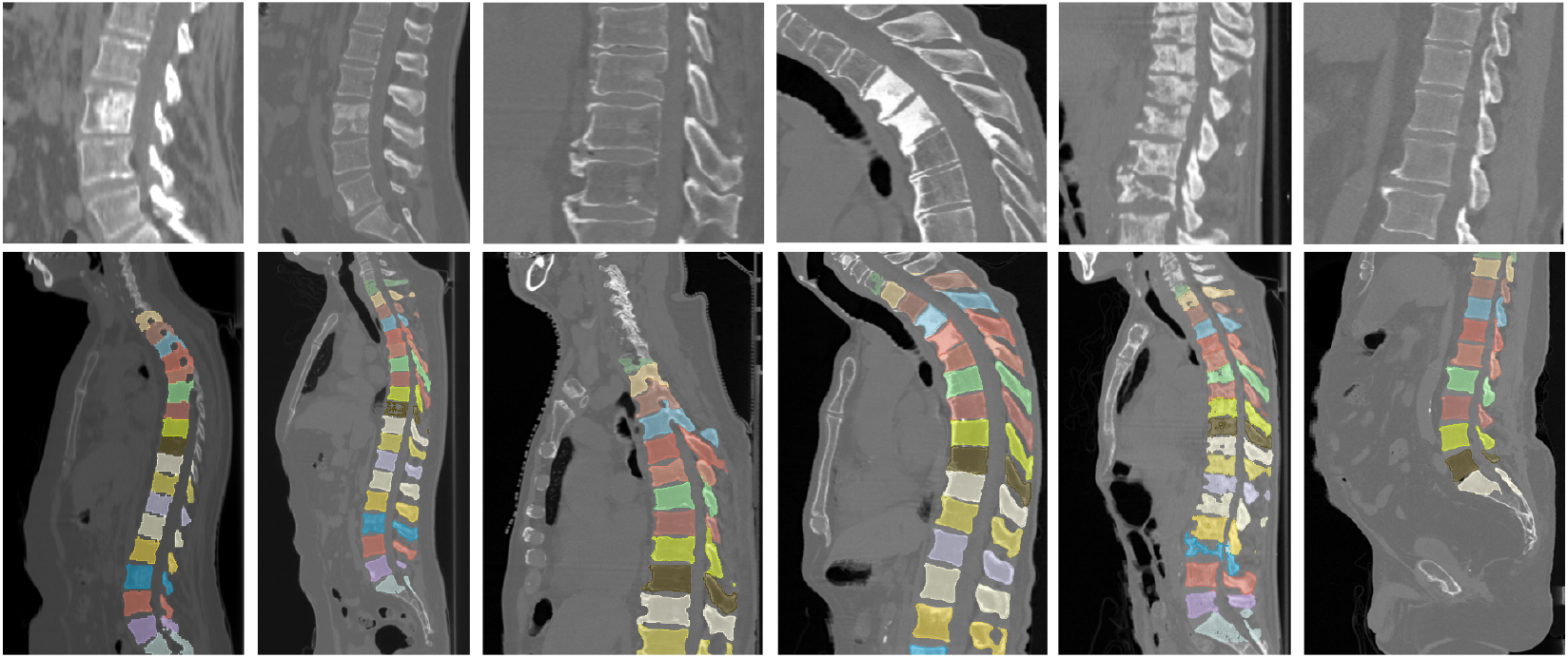
Example lesions and segmentations found in the dataset. Top row: Spine CTs with lytic, blastic, and mixed lesions and without lesions. Bottom row: Spine CTs with masks visualized as colored overlays.

**Figure 2:**
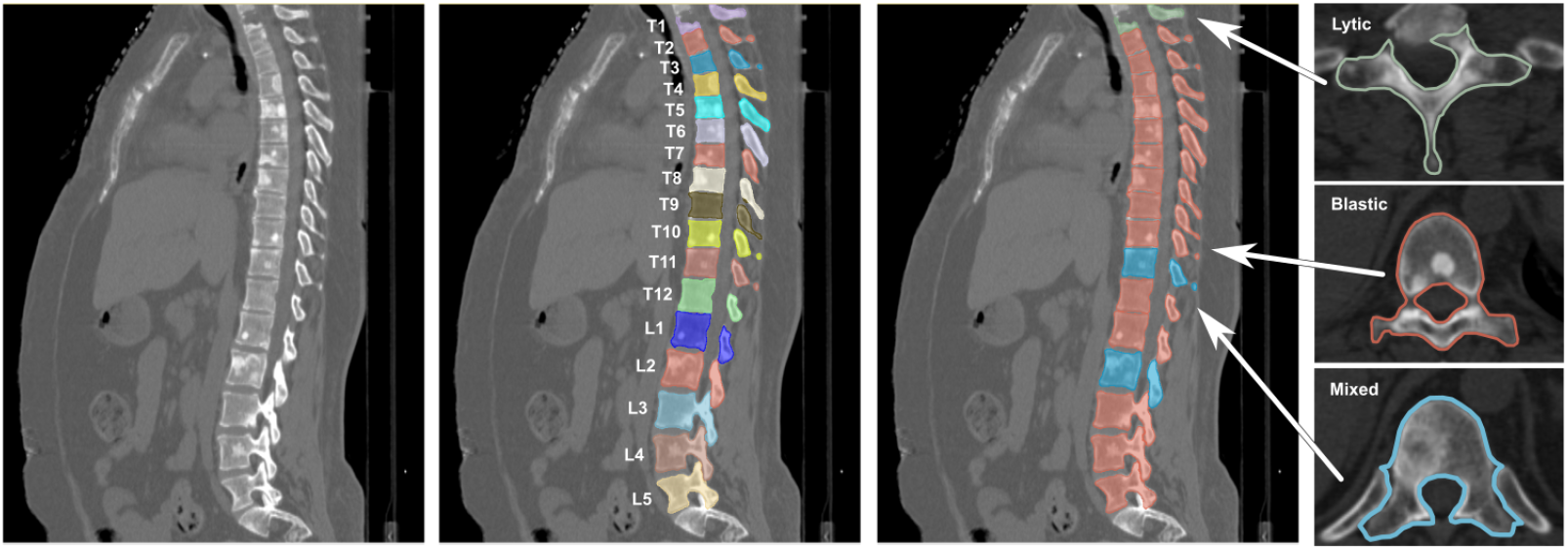
Illustrative example dataset of a patient with vertebral lesions with the raw CT, annotated segmentation and identification of vertebral levels, and lesion classification (Lytic, Blastic, and Mixed).

### Technical Validation: nnU-Net for Baseline Segmentation Performance

We trained a deep convolutional neural network using the dataset. Each vertebra was automatically segmented from the background, localized, and labeled, and 3D surfaces were generated using 3DSlicer. We used the nnU-Net framework (25), a self-adapting framework for 3D full-resolution image segmentation based on the U-Net architecture. We obtained an average segmentation accuracy of 94.33% ±2.24% Dice score on the spine CTs dataset not used in the training (3-fold validation). Segmentation results are illustrated in Figure 3.

**Figure 3:**
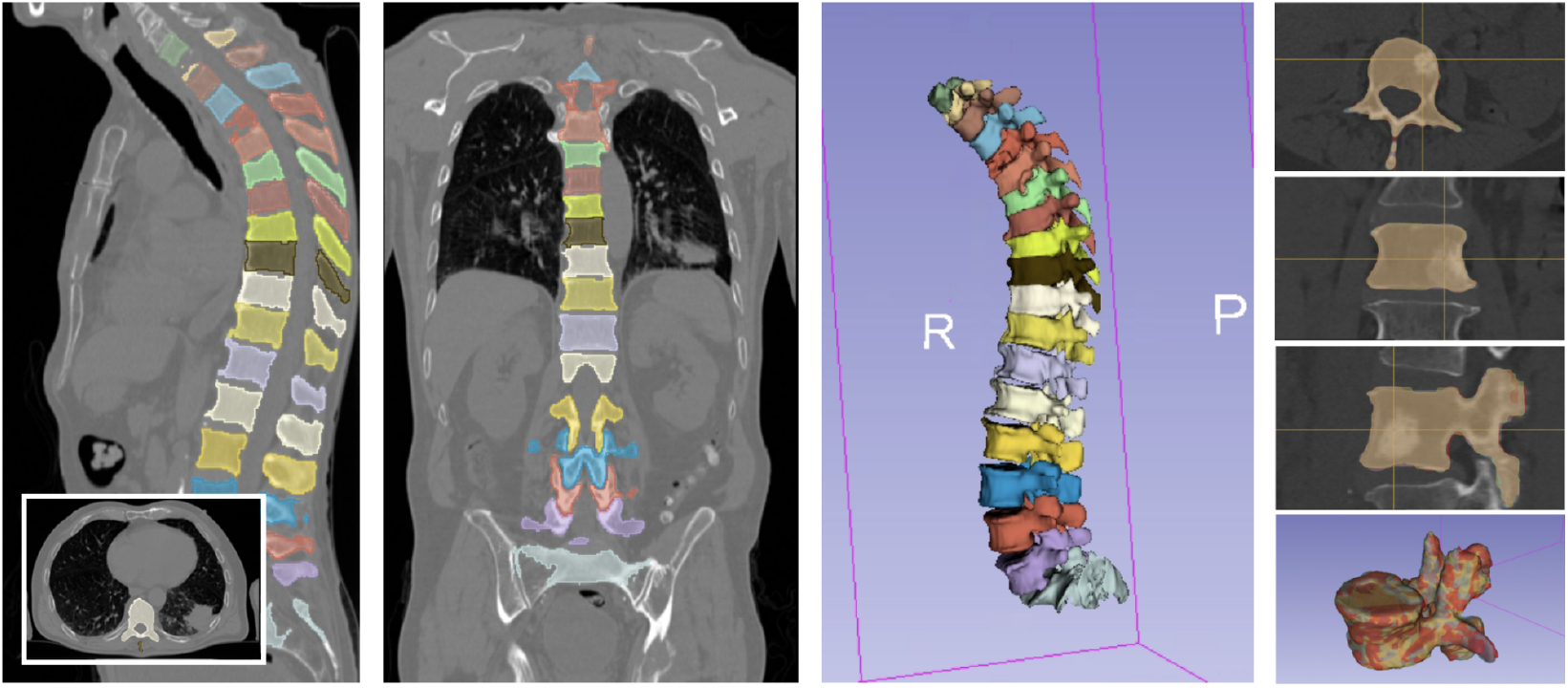
Segmentation results on one patient using the trained nnU-Net model. From left to right: Axial, Coronal, Sagittal, and 3D views of the spine CT with segmentation masks; segmentation results overlaid with ground truth annotation of one vertebra with a blastic lesion.

We then compared our model with the TotalSegmentator framework (15), trained on a spine CT dataset that excludes vertebral lesions. Our comparison exhibited an average improvement in Dice score of 3.32% across all spines and an average improvement in Dice score of 5.18% in vertebrae with bone metastasis, further validating the importance of our dataset.

## Resulting Dataset

De-identified data was submitted to The Cancer Imaging Archive (TCIA) through its established selection and governance processes. The CT images exclude the head and, thus, any facial features. The images and associated files are publicly available at:

**https://doi.org/10.7937/kh36-ds04**.

Both CT image files and segmentation are provided in DICOM format for each case, compatible with TCIA requirements and interoperating with PACS and commercial tools. We followed the SNOMED® CT coding scheme as the controlled terminology to label each vertebra. Lesion classification is provided in a Microsoft Excel format, and maps every vertebra level for each scan with a lesion class is also provided. In addition to the CT images, the dataset contains patient demographics data and primary cancer sites.

The dataset comprises 55 CT image series from 55 patients and 787 vertebrae, encompassing 543 thoracic and 244 lumbar vertebrae. Among these vertebrae, 183 contain lesions, 111 thoracic and 72 lumbar, classified as 58 blastic, 54 lytic, and 71 mixed (see Table 2). This represents a considerable increase in available annotated data—particularly for lesion vertebrae—compared with previously available datasets (17,18,19,20,21,22,26).

**Table 2:** Classification of vertebral lesions by level.

| Classification (Lesion) | Thoracic | Lumbar | Total |
| --- | --- | --- | --- |
| **Vertebra with Lesion** | 111 | 72 | 183 |
| - Blastic | 33 | 25 | 58 |
| - Lytic | 30 | 24 | 54 |
| - Mixed | 48 | 23 | 71 |
| **Vertebra Not Involved** | 432 | 172 | 604 |
| **All vertebrae** | 543 | 244 | 787 |

## Discussion

We present a unique dataset of spine CTs featuring metastatic bone lesions with annotated vertebral segmentation and lesion classifications. While public datasets of spine CTs, such as VerSe, are available, they primarily focus on healthy subjects and are dedicated to voxel-based segmentation and classification. With this dataset, we aim to enhance existing resources by specifically highlighting bone lesions to encourage further research in this domain.

We acknowledge that the dataset includes a limited number of cases, largely due to the complexity of lesion type classification, and does not contain follow-up scans. Nevertheless, we are confident that this resource will advance research in deep-learning approaches for detecting, segmenting and classifying metastatic bone lesions in spine CT. Ultimately, this dataset may support the development of objective quantitative criteria to guide treatment decisions aimed at reducing fracture risk and controlling pain in affected patients.

## Data Availability

The images and associated files are publicly available on The Cancer Imaging Archive.

https://doi.org/10.7937/kh36-ds04

## Acknowledgments

This work was supported by the NIAMS and the NIBIB of the National Institutes of Health under award numbers R01AR075964, 3R01AR075964-03S1, and K25EB035166.

## Notes

### Competing Interest Statement

The authors have declared no competing interest.

### Author Declarations

Ethics committee/IRB or Beth Israel Deaconess Medical Center, Brigham and Women's Hospital and Dana Farber Cancer Institute gave ethical approval for this work.

### Summary of Updates

Added two tables on demographics and details on the dataset. Shortened some sections for clarity.

